# Determinants of SARS-CoV-2 IgG response and decay in Canadian healthcare workers: a prospective cohort study

**DOI:** 10.1101/2023.09.12.23295445

**Authors:** Nicola Cherry, Anil Adisesh, Igor Burstyn, Carmen Charlton, Yan Chen, Quentin Durand-Moreau, France Labrèche, Shannon Ruzycki, LeeAnn Turnbull, Tanis Zadunayski, Yutaka Yasui

## Abstract

**Introduction:** Healthcare workers (HCWs) from an interprovincial Canadian cohort were asked to give serial blood samples to identify factors associated with anti-receptor binding domain (anti-RBD) IgG response to the SARS-CoV-2 virus.

**Methods:** Members of the HCW cohort donated blood samples four months after their first SARS-CoV-2 immunization and again at 7, 10 and 13 months. Date and type of immunizations and dates of SARS-CoV-2 infection were collected at each of four contacts, together with information on immunologically-compromising conditions and current therapies. Blood samples were analyzed centrally for anti-RBD IgG and anti-nucleocapsid IgG (Abbott Architect, Abbott Diagnostics). Records of immunization and SARS-CoV-2 testing from public health agencies were used to assess the impact of reporting errors on estimates from the random-effects multivariable model fitted to the data.

**Results:** 2752 of 4567 vaccinated cohort participants agreed to donate at least one blood sample. Modelling of anti-RBD IgG titer from 8903 samples showed an increase in IgG with each vaccine dose and with first infection. A decrease in IgG titer was found with the number of months since vaccination or infection, with the sharpest decline after the third dose. An immunization regime that included mRNA1273 (Moderna) resulted in higher anti-RBD IgG. Participants reporting multiple sclerosis, rheumatoid arthritis or taking selective immunosuppressants, tumor necrosis factor inhibitors, calcineurin inhibitors and antineoplastic agents had lower anti-RBD IgG. Supplementary analyses showed higher anti-RBD IgG in those reporting side-effects of vaccination, no relation of anti-RBD IgG to obesity and lower titers in women immunized early in pregnancy. Sensitivity analysis results suggested no important bias in the self-report data.

**Conclusion:** Creation of a prospective cohort was central to the credibility of results presented here. Serial serology assessments, with longitudinal analysis, provided effect estimates with enhanced accuracy and a clearer understanding of medical and other factors affecting response to vaccination.

## Introduction

In the early weeks of the COVID-19 pandemic, a cohort of healthcare workers (HCWs) was established in four Canadian provinces to determine modifiable workplace factors associated with increased risk of infection and of mental distress when working through the pandemic. Although this project envisaged using serology samples to identify or confirm SARS-CoV-2 infection, the need for serial samples of asymptomatic HCWs only became apparent with the start of SARS-CoV-2 immunization in December 2020. In response to a call from the Canadian Immunology Task Force for data to better understand the immunological response to SARS-CoV-2 vaccines, we established a serology sub-cohort to collect serial blood samples for determination of SARS-CoV-2 antibodies. At that point in the pandemic there was little knowledge about the effectiveness of vaccinations or the decay in immunological response, although this rapidly became available from clinical trials [1][2][3] and observational studies [4][5]6] [7]. Much useful data has come from cohort studies of HCWs [8] [9] [10] [11] [12] that were set up close to the start of vaccination against COVID-19, but long-term follow-up with repeated measures beyond the first months of vaccination are less in evidence [12] [13]. Using data from a serology sub-cohort of Canadian HCWs, we sought to establish an overall model of the relation between anti-receptor binding domain (anti-RBD) IgG to the spike protein of the SARS-CoV-2 virus (anti-RBD IgG titer) and the pattern of SARS-CoV-2 vaccinations and infections over time. As the serology sub-study was embedded in an on-going cohort, we were able to also consider wider questions raised elsewhere, including the impact of immune-related medical condition [14] [15] and medications [16] [17], the role, if any, of obesity [18] and pregnancy [19] and the relation of reported side-effects of vaccination to the strength of the anti-RBD IgG response [20]. Since the start of serology sampling in this cohort, knowledge on the immunological response to vaccines and infection and the decay over time has increased exponentially [21]. The results from the statistical models developed here to address these questions can be interpreted in this vastly better-informed context.

## Methods

The initial cohort recruited HCWs from four Canadian provinces (Alberta, British Columbia, Ontario and Quebec) in the early months (April-October) of the 2020 COVID-19 pandemic. This is described more fully elsewhere [22] but, in brief, HCWs were approached through their provincial professional organizations and invited to contact the research team if they were interested to take part. Physicians were recruited from all four provinces, registered and licensed practical nurses and health care aides only from Alberta and personal support workers only from Ontario. All consenting participants completed an online baseline/recruitment questionnaire in Spring/Summer 2020 and were contacted to complete further online questionnaires in late fall 2020, in April 2021 and April 2022. The second questionnaire asked if the HCW would donate a blood sample to look for undiagnosed COVID-19 via detection of anti-nucleocapsid IgG, emphasized a request to notify the research team of all positive tests and requested consent to obtain records of positive SARS-CoV-2 diagnostic nucleic acid tests (NAT) including polymerase chain reaction (PCR) tests from the public health authorities of their home province. The 3^rd^ and 4^th^ questionnaires asked for details of all vaccinations received and, in the 4^th^ questionnaire, consent to obtain immunization records from the province. Shortly after the first immunizations (19^th^ December 2020) participants were told about the plan to collect serial serology blood samples from those who were willing and who felt it was feasible. Samples were collected 4, 7, 10 and 13 months after the first vaccine dose. At the time of each sample, the participant was asked to complete a very brief questionnaire (online or by phone) to update their record of immunizations (date and manufacturer) and infection history, with details of how this was confirmed (by PCR (NAT) or rapid antigen testing).

Blood samples were collected through commercial or provincial blood collection facilities. These covered the whole province of Alberta but only parts of other provinces. In British Columbia (BC), with the support of the BC Centre for Disease Control, those outside the area covered by the commercial service could give samples through the BC public clinical laboratories. In Ontario and Quebec, those not covered by the commercial services could only give samples through ad hoc arrangements. Some did so, but for many it was not feasible to give repeated samples. All samples were forwarded to the Alberta BioSample Repository at the University of Alberta and stored at –20C for later analysis. This was carried out using the Abbott Architect platform (Abbott Diagnostics, Chicago, IL, USA) with the SARS-CoV-2 IgG II QUANT assay quantitatively detecting antibodies directed against the RBD region of the spike protein. The result of this assay, referred to as the anti-RBD IgG titer, is the outcome measure used throughout this article. The quantifiable range was 6.8-40,000 AU/ml: a value of ≥50 AU/ml was interpreted as a positive result. Samples were analyzed as they were received, at either of two laboratory centers, to give timely feedback to participants. Any participant whose sample result was <50AU/ml was contacted personally by the Principal Investigator to discuss likely causes and implications. To minimize the possibility of changes over time (in laboratory reagents, machines or procedures) that could lead to biased results in data serially analyzed, all samples with sufficient residual serum were reanalyzed for anti-RBD IgG under carefully standardized conditions in a single laboratory during summer of 2022. The results of the standardized reanalysis have been used for the present analysis, when available. Where no serum remained, the original result was retained. The Abbott Architect SARS-CoV-2 IgG antibody assay was also used to detect antibodies against the viral nucleocapsid protein, reflecting infection with the SARS-CoV-2 virus. In line with the manufacturer’s recommendation, a nucleocapsid result was considered positive with a value of 1.4 AU/ml or greater. The time between each vaccine dose and the date of sample collection was calculated for each of the serology samples. Vaccination occurring less than seven days prior to sample collection was not considered for that sample. The date of infection with COVID-19 was taken as the reported date of a positive PCR (NAT) or rapid antigen test.

Immunological conditions were ascertained from a checklist of six conditions plus ‘other’ and ‘none’ on the third and fourth questionnaire (Supplementary materials 11). Medications assessed were those reported by participants as likely or possibly affecting response to the vaccine (Supplementary materials 1.2). As there were few changes to medication over the period of vaccinations, those started before the first vaccine were used in the analysis. Responses to the checklist of autoimmune conditions (rheumatoid arthritis, ankylosing spondylitis, psoriatic arthritis, psoriasis, inflammatory bowel disease, and lupus) were accepted without review. Three physician members of the team reviewed all free-text entries entered as “other chronic autoimmune inflammatory diseases” or volunteered in the medication section: multiple sclerosis was identified from these responses. Medications were coded into seven categories based on the international classification of diseases (ICD) 11^th^ revision [23] (World Health Organization, 2020): methotrexate, tumor necrosis factor (TNF) inhibitors, glucocorticoids, interleukin inhibitors, calcineurin inhibitors, selective immunosuppressants, and antineoplastic agents (Supplementary materials 1.3). Coding was done independently with disagreements solved by consensus.

Gender as self-reported on the recruitment questionnaire and age at each sample, calculated from date of birth, were included in each analysis.

Variables explored in the supplementary analyses were defined as follows. Body Mass Index (BMI) was calculated from self-report of height and weight as kg/m^3^, collected at the third contact. For pregnancy, date of each conception was collected at the 4^th^ contact. Pregnancy gestation at the time of vaccination could not be calculated exactly, but was estimated from the month of conception reported by the participant, with ‘early’ gestation including those with the vaccine dose in the same month as conception or in the two following months, ‘mid’ gestation in months 4 or 5 and ‘late’ gestation in months 6-10 of a continuing pregnancy. Adverse side effects at first and second vaccination dose were collected from a list of thirteen possible effects at each dose (Supplementary material 1.4). The participant was asked to mark on a visual analogue scale (from ‘not at all’ to ‘very’) how much they had been bothered by each symptom.

Two provinces (Alberta and British Columbia) were able to provide public health records of vaccination and infection for participants who consented. The self-report and public health records were reconciled by adding unreported vaccinations and correcting dates and types of vaccine. Any PCR(NAT) confirmed cases of COVID-19 from public health records were added and existing dates corrected. The reconciled records were used for the analysis reported here, with a sensitivity analysis comparing models from the original and reconciled data.

## Statistical methods

To reduce skewness, the natural logarithm of anti-RBD IgG concentration was used in the analysis. Mean and standard deviation (SD) of the log anti-RBD IgG were calculated for each category of factors and tabulated, along with the corresponding geometric mean of the original anti-RBD IgG concentration. Mean time in months and its SD from each vaccine dose, or confirmed case of COVID-19, to the collection of blood sample was calculated and tabulated for each of the 4 time points of blood sample collection and overall. We used a linear mixed model with a Gaussian random-intercept for each HCW to describe the variation of log anti-RBD IgG values across the HCWs by age and gender and how the log IgG levels changed over time according to the following factors: the presence/absence of the first and second confirmed COVID-19 infections; time from each infection to the IgG measurement; nucleocapsid positivity status (positive, negative, missing); reception of up to four SARS-CoV-2 vaccinations prior to the sample; time from each vaccination to collection of the sample; type of the vaccines (only Pfizer, any Moderna, and any non-mRNA vaccines). The times from the 1^st^ vaccination and the 2^nd^ vaccination were almost redundant (i.e., Pearson correlation = 0.95) and thus only the latter was entered to the models. The value ascribed to vaccine 1 (present for all participants) is the model intercept. To this initial model, we added the seven indicators of the medical conditions (multiple sclerosis, rheumatoid arthritis, ankylosing spondylitis, psoriatic arthritis, psoriasis, inflammatory bowel disease, and lupus) and seven indicators of medication use (methotrexate, TNF inhibitors, glucocorticoids, interleukin inhibitors, calcineurin inhibitors, selective immunosuppressants, and antineoplastic agents) to evaluate the association of these clinical factors with log anti-RBD IgG levels, with an indicator of missing medical data. Three supplementary analyses were conducted to assess the associations of the severity of adverse effects (on the 2^nd^ dose of vaccination), pregnancy at the time of vaccination, and BMI with the log anti-RBD IgG levels: this was accomplished by adding each of the three variables to the second model above. For the analysis of adverse reactions to vaccination a separate principal component analysis of log-transformed visual analogue scales was carried out for adverse responses ascribed to the 1^st^ and 2^nd^ doses. Unrotated components with eigen values >1.0 were extracted and scores retained for analysis against anti-RBD IgG. As a sensitivity analysis, the initial model was re-fit using only 2031 participants with 6630 samples (74% of the total) in Alberta and British Columbia who had been matched to provincial records of PCR (NAT) confirmed SARS-CoV-2 infection and vaccination. The Akaike Information Criterion (AIC) was calculated for each dataset to determine whether the fit improved when public health data were incorporated

All statistical analyses were conducted by Stata version 18 and SAS 9.4. Probabilities quoted were two-sided.

## Results

Of the 4964 HCWs consenting to join the prospective cohort, 2752 gave at least one post vaccine serology sample. This represented 60% (2752/4567) of those eligible, excluding 397 HCW not known to have been vaccinated (Figure 1). Of the 2752 giving samples, 292 gave 1, 323 two, 583 three and 1554 four, for a total of 8903 samples. Of these, 96.9% (8629/8903) had IgG values from the controlled reanalysis. The geometric mean (GM) of anti-RBD IgG decreased from the 4-month sample (2689 AU/ml) to 1190 AU/ml at 7 months then increased to 3091AU/ml at the 10-month sample and again to 10,282 AU/ml at 13 months. The GMs of anti-RBD IgG by month from March/April 2021, the first date for a 4-month sample since first vaccine, to July 2022, the end of sample collection, are shown in Supplementary materials 2. The lowest GM was in November 2021.

**Figure 1.**
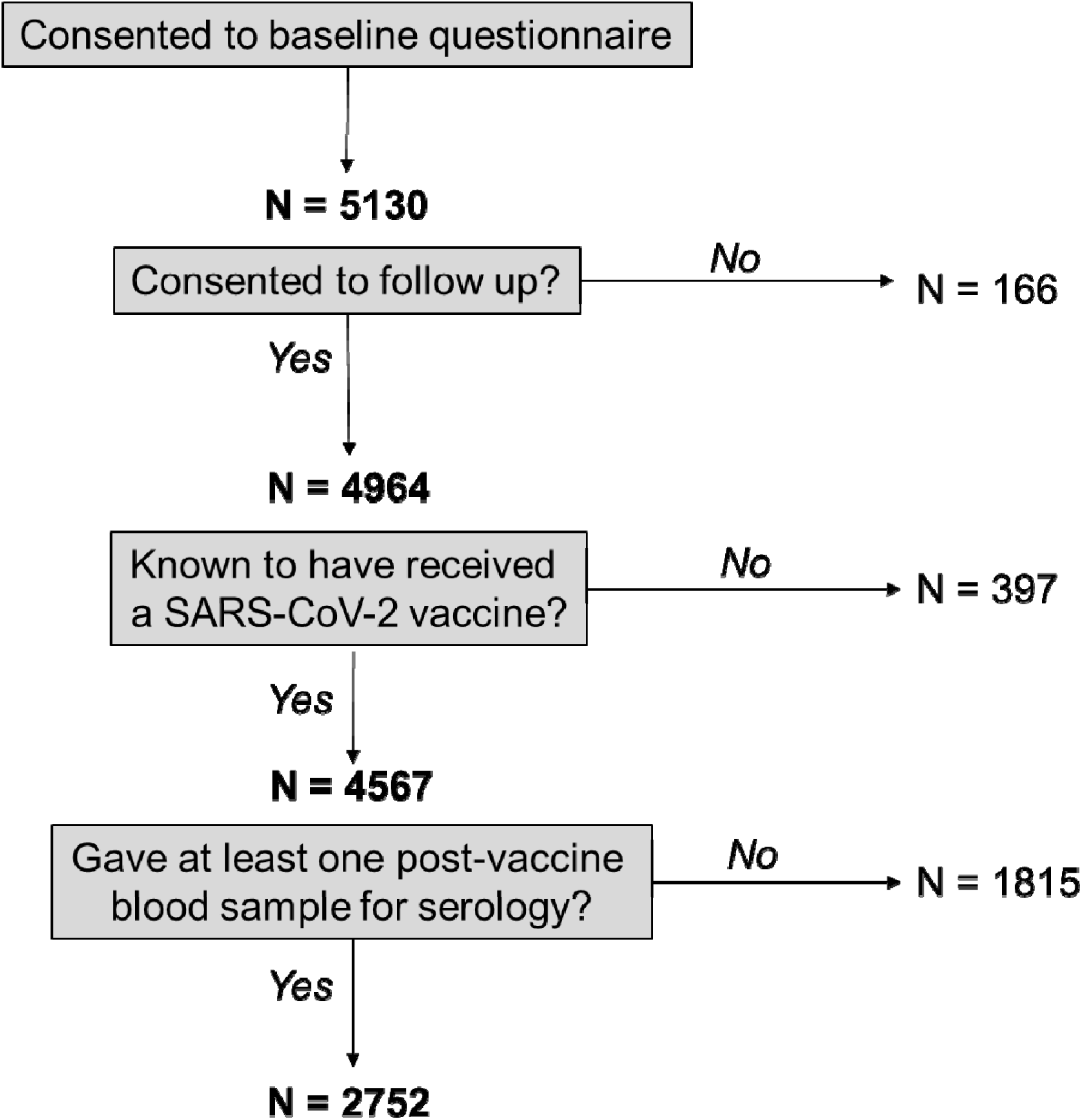
Flow chart of recruitment to the serology sub-study

The distribution of factors considered in the analysis is given in Table 1, together with the untransformed GM and mean log anti-RBD IgG unadjusted for other factors. The mean time between vaccines and infection prior to each sample is shown in Table 2. All but 46 of the 2216 participants giving a 4-month sample had received both first and second doses before this first sample. The initial model including gender, age and history of vaccination and infection is shown in Table 3. Gender was unrelated to the measured immune response and a decreasing response with age was only marked in those aged 65 or older. A positive effect on anti-RBD IgG was seen for each of four vaccine doses. Figure 2 shows the predicted values and slopes from the initial model (Table 3) for those who had received each of the four vaccines hypothetically at its mean time point of the cohort members (red line), together with smoothed observed values for those with (solid black) and without (dashed black) the third vaccine dose by the 10-month sample. The sharp increases in the predicted line reflect the estimated effects of the 3^rd^ and 4^th^ vaccine doses. The predicted slope post vaccine 2 is shallower (estimated –0.156) than that from vaccine 3 (–0.156 + –0.105 =-0.261) or vaccine 4 (–0.156 + –0.105 + 0.012= –0.249). The smoothed observed values follow the predicted line closely to the 7-month sample, diverge for the 10-month sample and meet again at the 13-month sample The predicted line in Figure 3 does not take account of infection or type of vaccine. A total of 678 of 2752 HCW in the serology sub-study had tested positive for SARS-CoV-2 by July 2022, of whom 19 had two test-positive infections (Table 1). The effect estimates for infection were positive, with the additional effect for the second case much smaller than the first (Table 3). The decay slope after infection was shallower than after vaccination. Having adjusted for known infection a positive nucleocapsid antibody result was related to a higher log anti-RBD IgG (Table 3). Those who had received only BNT162b2 (Pfizer-BioNTech) vaccines had lower anti-RBD IgG titers than those who had received at least one dose of mRNA-1273 (Moderna), while those whose doses included any non-mRNA vaccine had lower anti-RBD IgG (Table 3).

**Figure 2.**
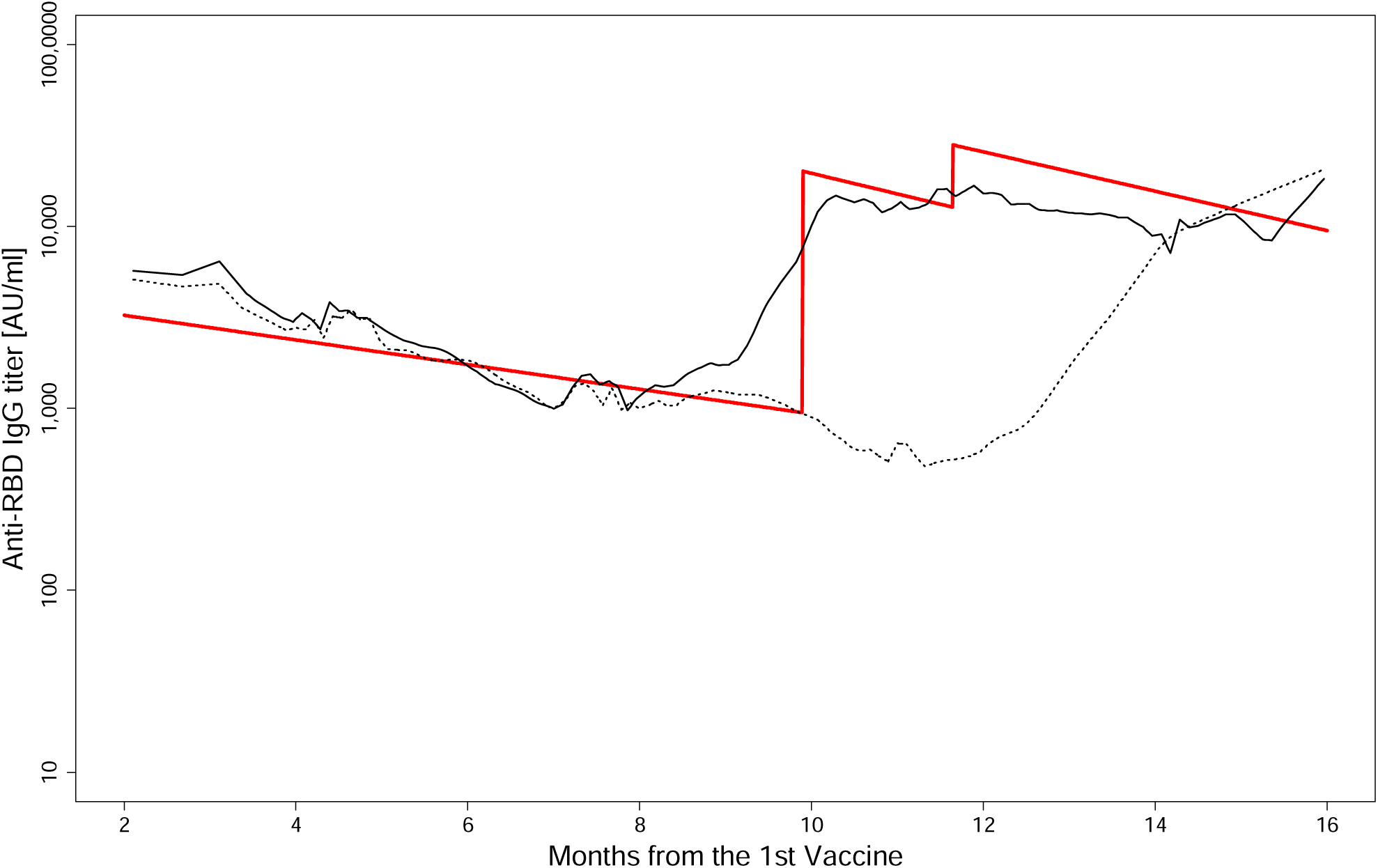
Anti-receptor binding domain (RBD) IgG titer to the spike protein of the SARS-CoV-2 virus by months from the 1^st^ vaccine The red line shows the model predicted anti-RBD IgG level for a person who is younger than 35 years old and received 4 vaccinations at the respective cohort-average time points of the 4 vaccinations. The vertical jumps around 10 and 12 months indicate the 3^rd^ and 4^th^ vaccine boosts of the IgG level, respectively. The solid and dotted black lines show the LOWESS (LOcally Weighted Scatterplot Smoothing) lines of observed log anti-RBD IgG levels for healthcare workers who had and had not, respectively, received the 3^rd^ vaccination before their 10-month samples. Note that the LOWESS lines do not take into account the longitudinal nature of the anti-RBD IgG levels within each healthcare worker across time points and just smooth the observed values of the respective groups.

**Table 1.** Geometric mean (GM) anti-RBD IgG and mean log anti-RBD IgG by personal characteristics, infection and vaccination at the time of sample collection.

| Factor |  | Anti-RBD IgG (GM)AU/ml | Log anti-RBD IgG |  | Number of |  |
| --- | --- | --- | --- | --- | --- | --- |
|  |  |  | Mean | SD | Observations | Participants |
| Gender | Female | 3249 | 8.09 | 1.56 | 7424 | 2292 |
|  | Other | 2977 | 8.00 | 1.54 | 1479 | 460 |
| Age (years) | < 35 | 3032 | 8.02 | 1.52 | 1291 | 439 |
|  | 35 < 50 | 3183 | 8.07 | 1.56 | 3472 | 1125 |
|  | 50 < 65 | 3317 | 8.11 | 1.56 | 3365 | 1065 |
|  | 65 or older | 3094 | 8.04 | 1.62 | 775 | 243 |
| COVID-19 | Confirmed 1<sup>st</sup> case | 12198 | 9.41 | 1.31 | 1127 | 678 |
|  | Confirmed 2<sup>nd</sup> case | 16248 | 9.70 | 0.78 | 26 | 19 |
| Nucleocapsid | Positive | 19583 | 9.88 | 1.11 | 668 | 544 |
|  | Missing | 2072 | 7.64 | 1.05 | 137 | 137 |
| Vaccination | Any 1<sup>st</sup> vaccine | 3202 | 8.07 | 1.56 | 8903 | 2752 |
|  | Any 2<sup>nd</sup> vaccine | 3246 | 8.09 | 1.55 | 8837 | 2737 |
|  | Any 3<sup>rd</sup> vaccine | 11175 | 9.32 | 1.10 | 3330 | 2222 |
|  | Any 4<sup>th</sup> vaccine | 9204 | 9.13 | 1.40 | 75 | 64 |
| Type of vaccine | Only Pfizer vaccine | 2740 | 7.92 | 1.56 | 7085 | 2263 |
|  | Any Moderna vaccine | 6323 | 8.75 | 1.39 | 1575 | 604 |
|  | Any non-mRNA vaccine | 3653 | 8.20 | 1.34 | 243 | 81 |
| Medical Condition | Multiple sclerosis | 279 | 5.63 | 2.84 | 43 | 12 |
|  | Rheumatoid arthritis | 1904 | 7.55 | 1.57 | 145 | 42 |
|  | Ankylosing spondylitis | 3513 | 8.16 | 1.56 | 48 | 16 |
|  | Psoriatic arthritis | 3368 | 8.12 | 1.65 | 54 | 18 |
|  | Psoriasis | 3026 | 8.02 | 1.65 | 271 | 88 |
|  | Inflammatory bowel disease | 3249 | 8.09 | 1.63 | 243 | 77 |
|  | Lupus | 2060 | 7.63 | 2.02 | 49 | 16 |
| Medication | Methotrexate | 1679 | 7.43 | 1.82 | 84 | 26 |
|  | Tumor Necrosis Factor inhibitor | 2098 | 7.65 | 1.67 | 61 | 21 |
|  | Glucocorticoids | 2163 | 7.68 | 1.69 | 25 | 8 |
|  | Interleukin inhibitor | 5056 | 8.53 | 1.20 | 23 | 7 |
|  | Calcineurin inhibitor | 702 | 6.55 | 2.45 | 11 | 3 |
|  | Selective immunosuppressants | 221 | 5.40 | 2.31 | 57 | 17 |
|  | Antineoplastic agents | 906 | 6.81 | 3.18 | 10 | 4 |
| Medical/medication | Data missing | 3709 | 8.22 | 1.44 | 49 | 18 |
| Body Mass Index | <25 | 3169 | 8.06 | 1.53 | 3992 | 1243 |
|  | 25<30 | 3162 | 8.06 | 1.55 | 2832 | 851 |
|  | 30<35 | 3161 | 8.06 | 1.57 | 1310 | 405 |
|  | 35<40 | 4042 | 8.30 | 1.73 | 388 | 121 |
|  | ≥40 | 3422 | 8.14 | 1.71 | 313 | 104 |
|  | Missing | 2510 | 7.83 | 1.49 | 68 | 28 |
| Vaccine dose during pregnancy | No | 3172 | 8.06 | 1.56 | 8689 | 2696 |
|  | Yes – early stage | 4429 | 8.40 | 1.67 | 97 | 42 |
|  | Yes – middle stage | 3840 | 8.25 | 1.55 | 82 | 35 |
|  | Yes – late stage | 6156 | 8.73 | 1.32 | 88 | 35 |
| Overall |  | 3202 | 8.07 | 1.56 | 8903 | 2752 |

**Table 2.** Mean time (months) between each vaccine dose or case of COVID-19 prior to collection of each blood sample.

|  | Nominal month of sample (since first vaccination). |  |  |  |  |  |  |  |  |  |  |  |  |  |  |
| --- | --- | --- | --- | --- | --- | --- | --- | --- | --- | --- | --- | --- | --- | --- | --- |
|  | 4-month sample |  |  | 7-month sample |  |  | 10-month sample |  |  | 13-month sample |  |  | Overall |  |  |
|  | Mean months | SD | N | Mean months | SD | N | Mean months | SD | N | Mean months | SD | N | Mean months | SD | N |
| Vaccine |  |  |  |  |  |  |  |  |  |  |  |  |  |  |  |
| 1 | 4.54 | 0.51 | 2216 | 7.87 | 0.53 | 2179 | 11.25 | 0.57 | 2258 | 14.51 | 0.68 | 2250 | 9.58 | 3.77 | 8903 |
| 2 | 2.90 | 0.87 | 2170 | 6.16 | 0.93 | 2170 | 9.54 | 1.02 | 2252 | 12.79 | 1.08 | 2245 | 7.91 | 3.82 | 8837 |
| 3 | 2.00 | 1.03 | 9 | 1.70 | 1.31 | 93 | 1.97 | 1.29 | 1146 | 4.17 | 1.66 | 2082 | 3.34 | 1.87 | 3330 |
| 4 | □ | □ | □ | 1.89 | □ | 1 | 2.25 | 1.26 | 13 | 2.57 | 1.89 | 61 | 2.50 | 1.78 | 75 |
| Case of COVID-19 |  |  |  |  |  |  |  |  |  |  |  |  |  |  |  |
| 1 | 7.93 | 5.27 | 9 | 8.79 | 5.90 | 15 | 6.17 | 6.57 | 50 | 5.15 | 6.28 | 604 | 5.34 | 6.30 | 678 |
| 2 | □ | □ | □ | □ | □ | □ | □ | □ | □ | 3.35 | 6.00 | 19 | 3.35 | 6.00 | 19 |

**Table 3.** Linear mixed-effects model of log anti-RBD IgG with vaccination and COVID-19 case history. (N=8903 samples from 2752 participants)

| Factor |  | Effect estimate | 95% CI |  | P = |
| --- | --- | --- | --- | --- | --- |
|  |  |  | Lower | Upper |  |
| Gender: | Female | 0.003 | -0.082 | 0.089 | 0.940 |
| Age (years) | <35 | 0.000 |  |  |  |
|  | 35<50 | -0.054 | -0.146 | 0.038 | 0.250 |
|  | 50<65 | -0.074 | -0.170 | 0.021 | 0.130 |
|  | 65 or greater | -0.234 | -0.366 | -0.102 | <0.001 |
| COVID-19 | Confirmed 1<sup>st</sup> case of COVID-19 | 1.388 | 1.280 | 1.495 | <0.001 |
|  | Months first case before sample | -0.049 | -0.061 | -0.037 | <0.001 |
|  | Confirmed 2<sup>nd</sup> case of COVID-19 | 0.084 | -0.380 | 0.547 | 0.720 |
|  | Months second case before sample | 0.005 | -0.067 | 0.078 | 0.890 |
|  | Nucleocapsid positive | 0.687 | 0.589 | 0.786 | <0.001 |
|  | Nucleocapsid data missing | -0.034 | -0.190 | 0.122 | 0.670 |
| Vaccination | Any first vaccine | 5.791 | 5.521 | 6.062 | <0.001 |
|  | Any second vaccine | 2.342 | 2.087 | 2.597 | <0.001 |
|  | Months second vaccine before sample | -0.156 | -0.164 | -0.148 | <0.001 |
|  | Any third vaccine | 3.056 | 2.983 | 3.128 | <0.001 |
|  | Months third vaccine before sample | -0.105 | -0.123 | -0.087 | <0.001 |
|  | Any fourth vaccine | 0.789 | 0.438 | 1.141 | <0.001 |
|  | Months fourth vaccine before sample | 0.012 | -0.106 | 0.130 | 0.840 |
| Type of vaccine | Only BNT162b2 (Pfizer/BioNTech) | 0.000 |  |  |  |
|  | Any mRNA1273 (Moderna) | 0.346 | 0.272 | 0.420 | <0.001 |
|  | Any non-mRNA vaccine | -0.456 | -0.632 | -0.281 | <0.001 |

The model was then extended to assess the effects of medical conditions and medications (Table 4). Among the medical conditions considered only HCWs with multiple sclerosis and rheumatoid arthritis had a lower than expected immune response, as indicated by anti-RBD IgG. Most of the medications considered were negatively related to the immune response but only tumor necrotic factor (TNF) inhibitors, calcineurin inhibitors, selective immunosuppressants and antineoplastic agents reduced the anti-RBD IgG concentration to a significant degree.

**Table 4.** Linear mixed-effects model of log anti-RBD IgG with medical conditions and medication. (N=8903 samples from 2752 participants)

| Factor |  | Effect estimate | 95% CI |  | P = |
| --- | --- | --- | --- | --- | --- |
|  |  |  | Lower | Upper |  |
| Gender: | Female | 0.031 | -0.051 | 0.113 | 0.46 |
| Age (years) | <35 | 0.000 |  |  |  |
|  | 35<50 | -0.068 | -0.157 | 0.021 | 0.13 |
|  | 50<65 | -0.074 | -0.166 | 0.017 | 0.11 |
|  | 65 or greater | -0.250 | -0.377 | -0.123 | <0.001 |
| Medical conditions | Multiple sclerosis | -1.619 | -2.178 | -1.060 | <0.001 |
|  | Rheumatoid arthritis | -0.383 | -0.672 | -0.093 | 0.010 |
|  | Ankylosing spondylitis | 0.388 | -0.040 | 0.817 | 0.076 |
|  | Psoriatic arthritis | -0.347 | -0.784 | 0.089 | 0.12 |
|  | Psoriasis | -0.102 | -0.281 | 0.076 | 0.26 |
|  | Inflammatory bowel disease | 0.028 | -0.164 | 0.220 | 0.78 |
|  | Lupus | 0.303 | -0.172 | 0.779 | 0.21 |
| Medication | Methotrexate | -0.025 | -0.429 | 0.378 | 0.90 |
|  | Tumor Necrosis Factor inhibitors | -0.712 | -1.111 | -0.313 | <0.001 |
|  | Glucocorticoid | -0.405 | -1.021 | 0.212 | 0.20 |
|  | Interleukin inhibitors | 0.419 | --0.202 | 1.041 | 0.19 |
|  | Calcineurin inhibitors | -1.149 | -2.047 | -0.251 | 0.012 |
|  | Selective immunosuppressants | -1.679 | -2.170 | -1.187 | <0.001 |
|  | Anti-neoplastic agents | -1.237 | -2.199 | -0.275 | 0.012 |
| Data missing on medical conditions/medication |  | 0.131 | -0.258 | 0.520 | 0.51 |
| COVID-19 | Confirmed 1<sup>st</sup> case of COVID-19 | 1.374 | 1.267 | 1.480 | <0.001 |
|  | Months first case before sample | -0.050 | -0.062 | -0.038 | <0.001 |
|  | Confirmed 2<sup>nd</sup> case of COVID-19 | 0.067 | -0.394 | 0.527 | 0.78 |
|  | Months second case before sample | 0.010 | -0.061 | 0.080 | 0.79 |
|  | Nucleocapsid positive | 0.691 | 0.593 | 0.789 | <0.001 |
|  | Nucleocapsid data missing | -0.038 | -0.193 | 0.117 | 0.63 |
| Vaccination | Any first vaccine | 5.836 | 5.570 | 6.102 | <0.001 |
|  | Any second vaccine | 2.319 | 2.067 | 2.571 | <0.001 |
|  | Months second vaccine before sample | -0.157 | -0.165 | -0.149 | <0.001 |
|  | Any third vaccine | 3.053 | 2.980 | 3.125 | <0.001 |
|  | Months third vaccine before sample | -0.101 | -0.120 | -0.083 | <0.001 |
|  | Any fourth vaccine | 0.838 | 0.487 | 1.189 | <0.001 |
|  | Months fourth vaccine before sample | 0.002 | -0.115 | 0.119 | 0.98 |
| Type of vaccine | Only BNT162b2 (Pfizer/BioNTech)) | 0.000 |  |  |  |
|  | Any mRNA1273 (Moderna) | 0.348 | 0.276 | 0.429 | <0.001 |
|  | Any non-mRNA vaccine | -0.487 | -0.656 | -0.317 | <0.001 |

Three supplementary analyses considered: whether severity of side effects to vaccination, gestation in women pregnant at the time of vaccination or body mass index (BMI), had any impact on the immune response, when these factors were added to the model shown in Table 4. To explore the relation of reported side effects to immune response (reflected in anti-RBD IgG concentration) three principal components analyses were extracted from responses to the 13 symptoms rated by respondents as their experience after the first and second vaccine dose (Table 5). Only the first components, interpreted as strength of systemic reactogenicity, related to an increased anti-RBD IgG response in bivariate analysis. The first components for the first and second vaccine doses were correlated (Pearson r= 0.53, p<0.001) but only the strength of response to the 2^nd^ dose added to the final model (Table 6). Of the 139 women in the serology sub-cohort who reported a pregnancy since the start of the pandemic, 88 received at least one vaccine dose during at least one pregnancy. These women appeared to have a somewhat lower response to vaccination if the vaccine dose was given in early or mid-pregnancy, when variables reflecting stage of pregnancy were added to the previous model (Table 6). With BMI in the model, there was no suggestion that participants with a higher BMI had a worse immune response (Table 6).

**Table 5.** Side effects reported for first and second vaccine doses. Mean log visual analogue scores with weights on the first 3 principal components extracted.

|  | First dose |  |  |  |  | Second dose |  |  |  |  |
| --- | --- | --- | --- | --- | --- | --- | --- | --- | --- | --- |
|  | Mean | SD | Component matrix* |  |  | Mean | SD | Component matrix* |  |  |
| Fatigue | 1.87 | 1.63 | 0.691 | -0.434 | 0.154 | 2.31 | 1.75 | 0.743 | -0.408 | 0.061 |
| Myalgia (muscle pain) | 1.62 | 1.60 | 0.685 | -0.335 | 0.080 | 1.84 | 1.73 | 0.777 | -0.345 | 0.001 |
| Arthralgia (joint pain) | 0.90 | 1.25 | 0.788 | -0.143 | -0.084 | 1.21 | 1.54 | 0.770 | -0.166 | -0.109 |
| Headache | 1.27 | 1.51 | 0.729 | -0.273 | 0.023 | 1.70 | 1.70 | 0.742 | -0.269 | 0.017 |
| Malaise | 1.19 | 1.48 | 0.796 | -0.322 | 0.024 | 1.76 | 1.74 | 0.788 | -0.381 | -0.040 |
| Feeling feverish | 0.80 | 1.20 | 0.811 | -0.089 | -0.168 | 0.78 | 1.17 | 0.517 | 0.352 | -0.224 |
| Chills | 0.77 | 1.19 | 0.802 | -0.048 | -0.179 | 1.28 | 1.60 | 0.753 | -0.179 | -0.105 |
| Diarrhea/loose stools | 0.60 | 0.96 | 0.701 | 0.346 | -0.219 | 0.66 | 1.02 | 0.650 | 0.379 | -0.250 |
| Nausea/vomiting | 0.62 | 0.99 | 0.753 | 0.252 | -0.227 | 0.77 | 1.16 | 0.680 | 0.247 | -0.207 |
| Skin reaction/rash | 0.55 | 0.87 | 0.646 | 0.499 | -0.083 | 0.57 | 0.89 | 0.611 | 0.547 | -0.045 |
| Swollen glands | 0.61 | 0.98 | 0.668 | 0.348 | -0.104 | 0.84 | 1.25 | 0.618 | 0.276 | -0.095 |
| Pain at injection site | 2.57 | 1.47 | 0.413 | 0.098 | 0.763 | 2.39 | 1.51 | 0.491 | 0.069 | 0.719 |
| Redness at injection site | 1.22 | 1.42 | 0.495 | 0.366 | 0.569 | 1.05 | 1.35 | 0.530 | 0.374 | 0.542 |
| N | 3989 |  |  |  |  | 2868 |  |  |  |  |

\*In the component analyses the variance accounted for by the first 3 components were:

|  | Component 1 | Component 2 | Component 3 |
| --- | --- | --- | --- |
| First vaccine dose | 49.0 | 9.3 | 8.6 |
| Second vaccine dose | 45.5 | 10.9 | 7.7 |

**Table 6.**
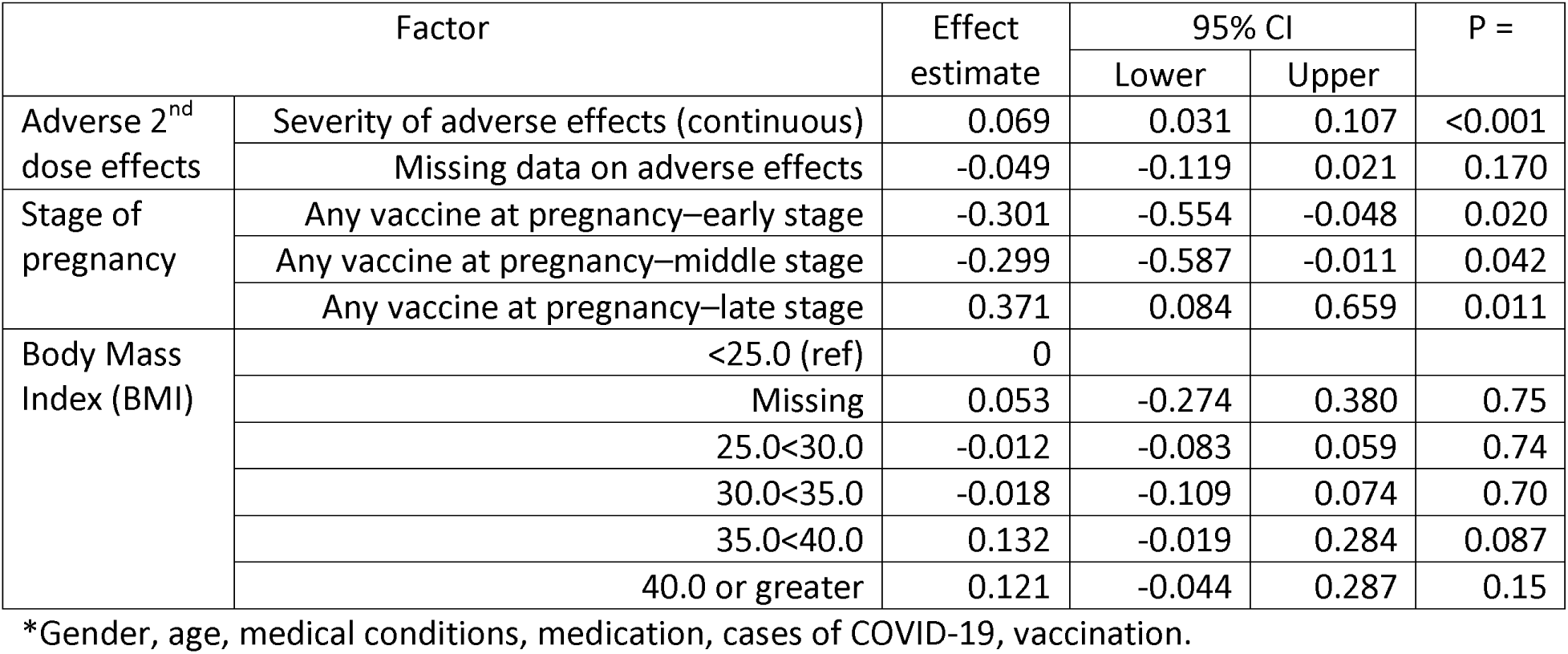
Relation of adverse effects to second vaccination, pregnancy and body mass index (BMI) to anti-RBD IgG adjusted for all factors* in Table 4.

In the sensitivity analysis (Table 7) estimates for 2031 participants with 6630 samples (74% of the total) in Alberta and British Columbia who had been matched to provincial public health records were very similar whether self-report alone or data reconciling official and self-report were used. The self-report data accounted for 75.8% of the variance in log anti-RBD IgG and the data corrected from public health records 79.8%. The AIC for the model with adjustment for public health data was smaller (19208) than with just the self-report (18220) suggesting the inclusion of public health records had improved the fit. Inspection of Table 7 indicates a greater (negative) effect of non-mRNA vaccines when self-reported vaccine type was corrected from public health records.

**Table 7.** Sensitivity analysis. Linear mixed effects models of log anti-RBD IgG for 2031 participants (6630 samples) for whom public health data were available.

| Factor |  | Self-report only |  |  |  | Self-report plus public health |  |  |  |
| --- | --- | --- | --- | --- | --- | --- | --- | --- | --- |
|  |  | Effect estimate | 95% CI |  | P= | Effect estimate | 95% CI |  | P= |
|  |  |  | Lower | Upper |  |  | Lower | Upper |  |
| Gender | Female | -0.029 | -0.137 | 0.080 | 0.600 | -0.035 | -0.138 | 0.069 | 0.510 |
| Age (years) | <35 | 0 |  |  |  | 0 |  |  |  |
|  | 35<50 | 0.019 | -0.091 | 0.129 | 0.730 | 0.024 | -0.080 | 0.128 | 0.650 |
|  | 50<65 | 0.002 | -0.111 | 0.115 | 0.970 | -0.004 | -0.112 | 0.103 | 0.940 |
|  | 65 or greater | -0.126 | -0.282 | 0.031 | 0.120 | -0.215 | -0.364 | -0.066 | 0.005 |
| COVID-19 | Confirmed 1<sup>st</sup> case of COVID-19 | 1.362 | 1.234 | 1.491 | <0.001 | 1.389 | 1.269 | 1.509 | <0.001 |
|  | Months first case before sample | -0.049 | -0.063 | -0.034 | <0.001 | -0.051 | -0.065 | -0.037 | <0.001 |
|  | Confirmed 2<sup>nd</sup> case of COVID-19 | -0.010 | -0.710 | 0.691 | 0.980 | -0.063 | -0.610 | 0.484 | 0.820 |
|  | Months second case before sample | -0.060 | -0.298 | 0.177 | 0.620 | 0.023 | -0.050 | 0.096 | 0.540 |
|  | Nucleocapsid positive | 0.736 | 0.619 | 0.853 | <0.001 | 0.685 | 0.576 | 0.794 | <0.001 |
|  | Nucleocapsid data missing | 0.090 | -0.090 | 0.271 | 0.330 | 0.033 | -0.133 | 0.200 | 0.700 |
| Vaccination | Any first vaccine | 6.209 | 5.890 | 6.529 | <0.001 | 5.166 | 4.795 | 5.538 | <0.001 |
|  | Any second vaccine | 1.745 | 1.446 | 2.045 | <0.001 | 2.952 | 2.593 | 3.312 | <0.001 |
|  | Months second vaccine before sample | -0.122 | -0.132 | -0.112 | <0.001 | -0.157 | -0.166 | -0.148 | <0.001 |
|  | Any third vaccine | 2.883 | 2.793 | 2.972 | <0.001 | 3.113 | 3.031 | 3.195 | <0.001 |
|  | Months third vaccine before sample | -0.120 | -0.142 | -0.098 | <0.001 | -0.108 | -0.129 | -0.088 | <0.001 |
|  | Any fourth vaccine | 0.722 | 0.263 | 1.181 | 0.002 | 0.739 | 0.324 | 1.154 | <0.001 |
|  | Months fourth vaccine before sample | -0.043 | -0.200 | 0.114 | 0.590 | 0.049 | -0.104 | 0.202 | 0.530 |
| Type of vaccine | Only BNT162b2 (Pfizer) | 0 |  |  |  | 0 |  |  |  |
|  | Any mRNA1273 (Moderna) | 0.308 | 0.221 | 0.395 | <0.001 | 0.296 | 0.215 | 0.377 | <0.001 |
|  | Any non-mRNA vaccine | -0.283 | -0.464 | -0.102 | 0.002 | -0.611 | -0.811 | -0.411 | <0.001 |

## Discussion

The serial collection of samples for serology testing demonstrated the anticipated increase of anti-RBD IgG with each vaccine dose and test-confirmed infection and the decrease in anti-RBD IgG titers with the passage of time. Further, it showed a greater increase in anti-RBD IgG with use of the mRNA-1273 vaccine, that nucleocapsid positivity was an independent predictor of anti-RBD IgG, the negative impact on anti-RBD IgG of certain medical conditions and treatments, a muted vaccine response in early pregnancy and an enhanced response in those reporting greater severity of common side effects.

The strengths of the study include the repeated measures design in a large cohort, with high participant retention, resulting in part from rapid feedback of serology results. A further strength is the very high proportion of samples included in the systematic reanalysis designed to minimize the impact of variability between laboratories and over time. The cohort design provided information on personal factors including medical conditions, adverse vaccine reactions, BMI and pregnancy. Although the information on vaccinations and infection was self-reported, the participants were all HCWs who may be more accurate in their reporting than the general population. Previous studies have shown that, in patients, self-report of vaccination status was very largely concordant with vaccination records but that self-reported dates and type of vaccine were less accurate [24] [25]. In the current study public health records were accessed for a majority of the serology sub-cohort and the congruence in estimates using purely self-report episodes and episodes corrected from external records gives some confidence to the findings for the serology cohort overall.

Weaknesses include the exclusion of a sizeable number of the main cohort for whom it was not feasible to obtain serial blood samples. Factors associated with joining the serology cohort have been described elsewhere [22]. In addition to those excluded through lack of local blood collection facilities, younger age, having a child at home and smoking were associated with a lower participation. The restricted serology analysis was a major limitation. Only serum was retained for analysis and it was not possible to infer anything about cellular immunity. While the sample size was adequate to confirm the negative impact of certain conditions and medications, it did not allow confident assertion of novel findings on the impact of pre-existing conditions. Moreover, this analysis does not consider the relation between vaccine response and breakthrough infection or severity of disease.

The use of 40,000 AU/ml as the upper limit of quantification produced the ceiling effect evident in Figure 2, and will have reduced the accuracy of estimates based on samples at 10 months (with 221 samples (9.8%) recorded at ceiling) and 13 months (425 samples, 18.9%). This will have biased the estimated slopes towards the null, as values potentially higher than 40,000 AU/ml were recorded as 40,000 AU/ml, resulting in estimates of decay slopes after the 3^rd^ and 4^th^ vaccine doses that are less steep than would have been expected from data without the 40000 AU/ml ceiling.

The observed increase in anti-RBD IgG following each vaccine dose and decay with time are as reported elsewhere [21] [26], as is the slower decay after infection in this cohort of vaccinated HCWs [21]. The smaller increase after the 4^th^ vaccine is consistent with other studies [13] but is based here on only 64 participants. Similarly, the small increase following a second infection was based on a confirmed second case in only 19 participants. The sharper decline after the 3^rd^ and 4^th^ vaccine dose than after the 2^nd^ is not consistent with some earlier reports which suggested a slower decline after the third dose [10] [12]. The reasons for this difference are uncertain. All three studies were of HCWs and all used the Abbott ARCHITECT system to obtain quantitative estimates of anti-RBD IgG. The current study was much larger and the follow-up after the third vaccine dose somewhat longer (mean 3.3 months) than in the previous studies cited here. Ikezaki et al [10] collected samples from 52 HCWs 2 months after the third vaccine dose and Dodge et al [12] from 212 HCWs 2-3 months after the third dose. The statistical approach differed between the studies. The analysis by Dodge et al [12] suggesting a shallower decay slope after the third vaccine considered only a subgroup in a cross-sectional analysis rather than a longitudinal analysis of IgG values which would have controlled for between-participant factors. Ikezaki et al [10] reported a percentage decline after the 2^nd^ and 3^rd^ vaccines without giving more detail. It should be noted that the effect of values above the upper limit of quantification, observed after the third but not second vaccine dose in all three studies, will have been differential, biasing estimates of decay after the third dose, but not the second, towards the null. The finding of a faster decline after the 3^rd^ vaccine dose in the current study is not easily dismissed: it is strengthened by the reanalysis of stored serum, ensuring results after the 2^nd^ and 3^rd^ doses were directly comparable. In their comprehensive review Sette and Crotty [21] commented that ‘conclusions about durability of [antibodies] after 3-dose mRNA vaccination remain uncertain’.

The relation of a positive nucleocapsid test, assayed from the same sample as the anti-RBD IgG, was related to a higher IgG concentration, whether or not there was a reported infection (data not shown). Without a reported case, this may be assumed to reflect an asymptomatic (or unrecognized) infection. In those with a recognized infection, this may again reflect a further unrecognized infection, or perhaps different rates of decay of anti-RBD IgG and nucleocapsid post infection [27]. The greater response in those with at least one dose of mRNA-1273 vaccine compared to those receiving only BNT162b2 is consistent with the literature [4] [12] [13] as is the lower IgG response with non-mRNA vaccines [7] [28] [29].

Our results show that HCWs with inflammatory medical conditions had lower levels of anti-RBD IgG, as reported elsewhere [30] [31]. Notably those participants with either multiple sclerosis or rheumatoid arthritis had a significantly lower concentration of anti-RBD IgG. These findings may be related to the condition itself affecting the immune response to vaccine or to the effects of medical treatment for the condition. When we examined the immune suppressant medications that participants reported, the lowest immune response was seen in those receiving one of the selective immunosuppressants (Supplementary materials 1.3). It has been shown that patients with multiple sclerosis (MS) taking these molecules have a decreased anti-spike protein IgG response to mRNA vaccines while untreated patients with MS had a response comparable to healthy controls [16]. Apostolidis et al [32] similarly demonstrated reduced anti-RBD IgG response to mRNA vaccines in a small group of patients with MS on anti-CD20 therapy. A recent registry study of patients with MS showed those treated with anti-CD20 therapies at the time of vaccination had “Abnormally Low” values [33]. In the current study, participants taking tumor necrosis factor-alpha inhibitors (TNFi) had significantly lower anti-RBD IgG as did the three participants prescribed calcineurin inhibitors. Kashiwado et al [17] reported that patients with auto-immune rheumatoid disease taking TNFi had lower anti-RBD IgG than healthy controls, with similar effects in patients taking TNFi for inflammatory bowel disease [34]. Only four participants reported taking antineoplastic agents at the time of their first vaccine dose. Their lower anti-RBD IgG response was consistent with that previously reported [35] but numbers were too low for analysis by agent.

Levy et al [20] reported on 831 HCWs for whom they collected adverse reactions to BNT162b2 mRNA vaccination through a questionnaire sent seven days after each vaccine dose. They examined the relation of reported adverse reactions to anti-RBD IgG in subsequent blood samples. As in the current study, they found that higher overall scores of systemic reactions to the second vaccine related to higher level of anti-RBD IgG. Two smaller studies of HCW from Japan [36] and South Korea [37] found no clear relation between anti-spike protein IgG and reported adverse events, while two from the United States [38] and Greece [9] again found those with common adverse reactions had greater anti-RBD IgG levels. In the current study the relation between adverse reactions after the second dose and anti-RBD IgG persisted after adjustment for many of the factors (including age, gender, vaccine type) that might act as confounders. Levy and colleagues [20] concluded that the mechanism of this association between immunological response and adverse effects was yet to be understood and that it was unclear whether there is causality in either direction: does a stronger immunological response produce more inflammatory mediators and products of inflammation that spill from the injection site to the general circulation, resulting in systemic symptoms [39] or does a systemic response trigger a more marked immunological one. Such questions have been raised also for pertussis [40] and adjuvants for the human papillomavirus [41]. The marked similarity of results between the Levy report and the current study suggests that there is indeed a phenomenon to be explained.

In the present study, in which the majority of participants were female, pregnancies during the pandemic were reported by many HCWs. While some chose to delay vaccination because of their pregnancy, sufficient had received at least one vaccine dose during pregnancy to allow us to explore earlier reports of lower anti-spike protein IgG [19] or anti-RBD IgG [42] after a vaccination dose received during pregnancy than in non-pregnant referents. Gray et al [43], looked at the same question and reported no effect although, graphically, anti-RBD IgG appeared lower in pregnant women post-vaccination. Atyeo et al [44] compared anti-spike protein IgG in 158 women receiving vaccines during pregnancy (11 first, 88 second and 52 third trimester) and found the highest (log) median IgG in the third trimester and lowest in the first trimester: this was reported to be ‘non-significant’ without further detail. The analysis reported here compared vaccine response in pregnant women to that of all healthcare workers. Although the information on gestation will have been less exact than in the previous studies from obstetric centers, the strength of the statistical modelling gives credence to the observation that reduction in antibody production (anti-RBD IgG) following vaccination was limited to early and mid-pregnancy.

There is limited evidence that obesity is a risk factor for a low IgG following vaccination. Watanabe et al [18] found that higher BMI in a sample of 86 HCWs related to lower anti-spike protein IgG in bivariate but not multivariate models. Waist circumference was more strongly related than BMI. Herzberg et al [45] reported that those with a BMI >25 had a lower anti-spike protein IgG response than others in their German HCW cohort. In a Greek cohort of HCWs, Papaioannidou et al [11] found, as in the present study, no clear evidence that BMI was a risk factor for a low anti-RBD IgG response. The interesting observation that neutralising antibodies were less effective and declined faster in those classified as severely obese [46], with no difference from controls in anti-RBD IgG [47], cannot be investigated further here. A systematic review [48] of the relation between humoral immunity and obesity reported a reduction in antibodies in people with obesity but did not differentiate between the types of antibodies measured.

The study reported here used serial blood samples, spaced over the first 18 months after the introduction of vaccines against SARS-CoV-2 in December 2020 and the end of the study in July 2022. By analysing these in a longitudinal model we have minimised the variance due to uncontrolled differences between participants. The resulting model confirms the now expected increase in anti-RBD IgG with successive vaccination and infections but identifies a faster decay after the third and fourth doses than after the second. Having established this model, we used it to determine whether there were other factors influencing this response. The results demonstrate the importance of recruiting and retaining cohorts with a rich body of observational data through which to better understand response to vaccination and to help identify areas in which questions still need to be answered.

## Criteria for authorship

All authors attest they meet the ICMJE criteria for authorship.

## Conflicts

The authors declare no conflict of interests.

## Ethics approval

Approval for each element of the study was given by the University of Alberta Heath Ethics Board (Pr000099700). The study was also reviewed and approved by Unity Health Toronto Research Ethics Board (REB# 20-298) for those elements coordinated locally for the Ontario participants. All participants gave online written informed consent after the nature and possible consequences of the study had been fully explained. The research was carried out in accordance with the Declaration of Helsinki.

## Author contributions

AA, IB, QDM, FL, SR and NC devised the project to establish the healthcare worker cohort and all actively collaborated to ensure the retention of participants and the serial collection of serology blood samples. LAT and CC took responsibility for the analysis of samples and resolution of disparities. TZ and YC curated the longitudinal dataset. YY, YC and NC carried out the data analysis. All authors contributed to drafting the paper, critically reviewed the final draft and approved the version submitted.

## Data availability statement

The data on which this manuscript is based will be made available on reasonable request to the corresponding author. The data are subject to a data sharing agreement with the major funder, the Canadian Immunology Task Force (CITF).. Through the databank currently being built individual anonymized data will be made available to researchers internationally on request to the CITF

## Acknowledgements

The analysis of serum samples was carried out by the Alberta Precision Laboratories and we are grateful to the Medical Director, Graham Tipples and to Paul Jacquier, Director, for their considerable help in ensuring the successful completion of the logistically complex analytical tasks. The data from public health records supplied through the Alberta and British Columbia health services were invaluable and made possible the sensitivity analysis. The interpretation and conclusions contained herein are those of the researchers and do not necessarily represent the views of the Governments of Alberta or British Columbia.

## References

1) Polack FP, Thomas SJ, Kitchin N, Absalon J, Gurtman A, Lockhart S, Perez JL, Pérez Marc G, Moreira ED, Zerbini C, Bailey R, Swanson KA, Roychoudhury S, Koury K, Li P, Kalina WV, Cooper D, Frenck RW Jr, Hammitt LL, Türeci Ö, Nell H, Schaefer A, Ünal S, Tresnan DB, Mather S, Dormitzer PR, Şahin U, Jansen KU, Gruber WC; C4591001 Clinical Trial Group. Safety and Efficacy of the BNT162b2 mRNA Covid-19 Vaccine. N Engl J Med. 2020 Dec 31;383(27):2603–2615. doi: 10.1056/NEJMoa2034577.

2) Baden LR, El Sahly HM, Essink B, Kotloff K, Frey S, Novak R, Diemert D, Spector SA, Rouphael N, Creech CB, McGettigan J, Khetan S, Segall N, Solis J, Brosz A, Fierro C, Schwartz H, Neuzil K, Corey L, Gilbert P, Janes H, Follmann D, Marovich M, Mascola J, Polakowski L, Ledgerwood J, Graham BS, Bennett H, Pajon R, Knightly C, Leav B, Deng W, Zhou H, Han S, Ivarsson M, Miller J, Zaks T; COVE Study Group. Efficacy and Safety of the mRNA-1273 SARS-CoV-2 Vaccine. N Engl J Med. 2021 Feb 4;384(5):403–416. doi: 10.1056/NEJMoa2035389.

3) Thomas SJ, Moreira ED Jr, Kitchin N, Absalon J, Gurtman A, Lockhart S, Perez JL, Pérez Marc G, Polack FP, Zerbini C, Bailey R, Swanson KA, Xu X, Roychoudhury S, Koury K, Bouguermouh S, Kalina WV, Cooper D, Frenck RW Jr, Hammitt LL, Türeci Ö, Nell H, Schaefer A, Ünal S, Yang Q, Liberator P, Tresnan DB, Mather S, Dormitzer PR, Şahin U, Gruber WC, Jansen KU; C4591001 Clinical Trial Group. Safety and Efficacy of the BNT162b2 mRNA Covid-19 Vaccine through 6 Months. N Engl J Med. 2021 Nov 4;385(19):1761–1773. doi: 10.1056/NEJMoa2110345.

4) Dickerman BA, Gerlovin H, Madenci AL, Kurgansky KE, Ferolito BR, Figueroa Muñiz MJ, Gagnon DR, Gaziano JM, Cho K, Casas JP, Hernán MA. Comparative Effectiveness of BNT162b2 and mRNA-1273 Vaccines in U.S. Veterans. N Engl J Med. 2022;386(2):105–115. doi:10.1056/NEJMoa2115463

5) Chemaitelly H, Tang P, Hasan MR, AlMukdad S, Yassine HM, Benslimane FM, Al Khatib HA, Coyle P, Ayoub HH, Al Kanaani Z, Al Kuwari E, Jeremijenko A, Kaleeckal AH, Latif AN, Shaik RM, Abdul Rahim HF, Nasrallah GK, Al Kuwari MG, Al Romaihi HE, Butt AA, Al-Thani MH, Al Khal A, Bertollini R, Abu-Raddad LJ. Waning of BNT162b2 Vaccine Protection against SARS-CoV-2 Infection in Qatar. N Engl J Med. 2021 Dec 9;385(24):e83. doi:10.1056/NEJMoa2114114.

6) Tartof SY, Slezak JM, Fischer H, Hong V, Ackerson BK, Ranasinghe ON, Frankland TB, Ogun OA, Zamparo JM, Gray S. Effectiveness of mRNA BNT162b2 COVID-19 vaccine up to 6 months in a large integrated health system in the USA: A retrospective cohort study. Lancet. 2021 Oct 16;398(10309):1407–1416. doi:10.1016/S0140-6736(21)02183-8.

7) Fedele G, Trentini F, Schiavoni I, Abrignani S, Antonelli G, Baldo V, Baldovin T, Bandera A, Bonura F, Clerici P, De Paschale M, Fortunato F, Gori A, Grifantini R, Icardi G, Lazzarotto T, Lodi V, Mastroianni CM, Orsi A, Prato R, Restivo V, Carsetti R, Piano Mortari E, Leone P, Olivetta E, Fiore S, Di Martino A, Brusaferro S, Merler S, Palamara AT, Stefanelli P. Evaluation of humoral and cellular response to four vaccines against COVID-19 in different age groups: A longitudinal study. Front Immunol. 2022 Oct 31;13:1021396. doi:10.3389/fimmu.2022.1021396.

8) Lustig Y, Sapir E, Regev-Yochay G, Cohen C, Fluss R, Olmer L, Indenbaum V, Mandelboim M, Doolman R, Amit S, Mendelson E, Ziv A, Huppert A, Rubin C, Freedman L, Kreiss Y. BNT162b2 COVID-19 vaccine and correlates of humoral immune responses and dynamics: a prospective, single-centre, longitudinal cohort study in health-care workers. Lancet Respir Med. 2021 Sep;9(9):999–1009. doi:10.1016/S2213-2600(21)00220-4.

9) Hatzakis A, Karabinis A, Roussos S, Pantazis N, Degiannis D, Chaidaroglou A, Petsios K, Pavlopoulou I, Tsiodras S, Paraskevis D, Sypsa V, Psichogiou M. Modelling SARS-CoV-2 Binding Antibody Waning 8 Months after BNT162b2 Vaccination. Vaccines (Basel). 2022 Feb 13;10(2):285. doi: 10.3390/vaccines10020285.

10) Ikezaki H, Nomura H, Shimono N. Dynamics of anti-spike IgG antibody after a third BNT162b2 COVID-19 vaccination in Japanese health care workers. Heliyon. 2022 Dec;8(12):e12125. doi: 10.1016/j.heliyon.2022.e12125.

11) Papaioannidou P, Skoumpa K, Bostanitis C, Michailidou M, Stergiopoulou T, Bostanitis I, Tsalidou M. Age, Sex and BMI Relations with Anti-SARS-CoV-2-Spike IgG Antibodies after BNT162b2 COVID-19 Vaccine in Health Care Workers in Northern Greece. Microorganisms. 2023 May 13;11(5):1279. doi: 10.3390/microorganisms11051279.

12) Dodge MC, Ye L, Duffy ER, Cole M, Gawel SH, Werler MM, Daghfal D, Andry C, Kataria Y. Kinetics of SARS-CoV-2 Serum Antibodies Through the Alpha, Delta, and Omicron Surges Among Vaccinated Health Care Workers at a Boston Hospital. Open Forum Infect Dis. 2023 May 17;10(7):ofad266. doi:10.1093/ofid/ofad266.

13) Canetti M, Barda N, Gilboa M, Indenbaum V, Mandelboim M, Gonen T, Asraf K, Weiss-Ottolenghi Y, Amit S, Doolman R, Mendelson E, Harats D, Freedman LS, Kreiss Y, Lustig Y, Regev-Yochay G. Immunogenicity and efficacy of fourth BNT162b2 and mRNA1273 COVID-19 vaccine doses; three months follow-up. Nat Commun. 2022 Dec 13;13(1):7711. doi:10.1038/s41467-022-35480-2.

14) Lee ARYB, Wong SY, Chai LYA, Lee SC, Lee MX, Muthiah MD, Tay SH, Teo CB, Tan BKJ, Chan YH, Sundar R, Soon YY. Efficacy of covid-19 vaccines in immunocompromised patients: systematic review and meta-analysis. BMJ. 2022 Mar 2;376:e068632. doi: 10.1136/bmj-2021-068632.

15) Al-Haideri M, Mohammad TAM, Darvishzadehdeldari S, Karbasi Z, Alimohammadi M, Faramarzi F, Khorasani S, Rasouli A, Tahmasebi S, Darvishi M, Akhavan-Sigari R. Immunogenicity of COVID-19 vaccines in adult patients with autoimmune inflammatory rheumatic diseases: A systematic review and meta-analysis. Int J Rheum Dis. 2023 Jul;26(7):1227–1234. doi: 10.1111/1756-185X.14713.

16) Achiron A, Mandel M, Dreyer-Alster S, Harari G, Magalashvili D, Sonis P, Dolev M, Menascu S, Flechter S, Falb R, Gurevich M. Humoral immune response to COVID-19 mRNA vaccine in patients with multiple sclerosis treated with high-efficacy disease-modifying therapies. Ther Adv Neurol Disord. 2021 Apr 22;14:17562864211012835. doi: 10.1177/17562864211012835.

17) Kashiwado Y, Kimoto Y, Ohshima S, Sawabe T, Irino K, Nakano S, Hiura J, Yonekawa A, Wang Q, Doi G, Ayano M, Mitoma H, Ono N, Arinobu Y, Niiro H, Hotta T, Kang D, Shimono N, Akashi K, Takeuchi T, Horiuchi T. Immunosuppressive therapy and humoral response to third mRNA COVID-19 vaccination with a six-month interval in rheumatic disease patients. Rheumatology (Oxford). 2023 Jun 8:kead275. doi: 10.1093/rheumatology/kead275.

18) Watanabe M, Balena A, Tuccinardi D, Tozzi R, Risi R, Masi D, Caputi A, Rossetti R, Spoltore ME, Filippi V, Gangitano E, Manfrini S, Mariani S, Lubrano C, Lenzi A, Mastroianni C, Gnessi L. Central obesity, smoking habit, and hypertension are associated with lower antibody titres in response to COVID-19 mRNA vaccine. Diabetes Metab Res Rev. 2022 Jan;38(1):e3465. doi:10.1002/dmrr.3465.

19) Blakeway H, Amin-Chowdhury Z, Prasad S, Kalafat E, Ismail M, Abdallah FN, Rezvani A, Amirthalingam G, Brown K, Le Doare K, Heath PT, Ladhani SN, Khalil A. Evaluation of immunogenicity and reactogenicity of COVID-19 vaccines in pregnant women. Ultrasound Obstet Gynecol. 2022 Nov;60(5):673–680. doi: 10.1002/uog.26050.

20) Levy I, Levin EG, Olmer L, Regev-Yochay G, Agmon-Levin N, Wieder-Finesod A, Indenbaum V, Herzog K, Doolman R, Asraf K, Halperin R, Lustig Y, Rahav G. Correlation between Adverse Events and Antibody Titers among Healthcare Workers Vaccinated with BNT162b2 mRNA COVID-19 Vaccine. Vaccines (Basel). 2022 Jul 30;10(8):1220. doi:10.3390/vaccines10081220.

21) Sette A, Crotty S. Immunological memory to SARS-CoV-2 infection and COVID-19 vaccines. Immunol Rev. 2022 Sep;310(1):27–46. doi: 10.1111/imr.13089.

22) Cherry N, Adisesh A, Burstyn I, Durand-Moreau Q, Galarneau J-M, Labrèche F, Ruzycki S, Zadunayski T. Cohort profile: Recruitment and retention in a prospective cohort of Canadian health care workers during the COVID-19 pandemic. Under review by BMJ Open and available as a preprint at 10.1101/2023.04.14.23288575 Accessed 26 April 2023.

23) World Health Organization (2020). International Classification of Diseases 11th Revision. Available on: https://icd.who.int/en (accessed on 11 August 2023).

24) Archambault PM, Rosychuk RJ, Audet M, Bola R, Vatanpour S, Brooks SC, Daoust R, Clark G, Grant L, Vaillancourt S, Welsford M, Morrison LJ, Hohl CM, Canadian COVID-19 Emergency Department Rapid Response Network (CCEDRRN) investigators, Network of Canadian Emergency Researchers, Canadian Critical Care Trials Group. Accuracy of Self-Reported COVID-19 Vaccination Status Compared With a Public Health Vaccination Registry in Québec: Observational Diagnostic Study. JMIR Public Health Surveill. 2023;9:e44465. doi:10.2196/44465.

25) Stephenson M, Olson SM, Self WH, Ginde AA, Mohr NM, Gaglani M, Shapiro NI, Gibbs KW, Hager DN, Prekker ME, Gong MN, Steingrub JS, Peltan ID, Martin ET, Reddy R, Busse LW, Duggal A, Wilson JG, Qadir N, Mallow C, Kwon JH, Exline MC, Chappell JD, Lauring AS, Baughman A, Lindsell CJ, Hart KW, Lewis NM, Patel MM, Tenforde MW, Network Investigators IV. Ascertainment of vaccination status by self-report versus source documentation: impact on measuring COVID-19 vaccine effectiveness. Influenza Other Respir Viruses. 2022 Nov;16(6):1101–1111. doi:10.1111/irv.13023.

26) Israel A, Shenhar Y, Green I, Merzon E, Golan-Cohen A, Schäffer AA, Ruppin E, Vinker S, Magen E. Large-Scale Study of Antibody Titer Decay following BNT162b2 mRNA Vaccine or SARS-CoV-2 Infection. Vaccines (Basel). 2021 Dec 31;10(1):64. doi: 10.3390/vaccines10010064.

27) Navaratnam AMD, Shrotri M, Nguyen V, Braithwaite I, Beale S, Byrne TE, Fong WLE, Fragaszy E, Geismar C, Hoskins S, Kovar J, Patel P, Yavlinsky A, Aryee A, Rodger A, Hayward AC, Aldridge RW; Virus Watch Collaborative. Nucleocapsid and spike antibody responses following virologically confirmed SARS-CoV-2 infection: an observational analysis in the Virus Watch community cohort. Int J Infect Dis. 2022 Oct;123:104–111. doi:10.1016/j.ijid.2022.07.053.

28) Steensels D, Pierlet N, Penders J, Mesotten D, Heylen L. Comparison of SARS-CoV-2 Antibody Response Following Vaccination With BNT162b2 and mRNA-1273. JAMA. 2021 Oct 19;326(15):1533–1535. doi:10.1001/jama.2021.15125.

29) Wei J, Pouwels KB, Stoesser N, Matthews PC, Diamond I, Studley R, Rourke E, Cook D, Bell JI, Newton JN, Farrar J, Howarth A, Marsden BD, Hoosdally S, Jones EY, Stuart DI, Crook DW, Peto TEA, Walker AS, Eyre DW; COVID-19 Infection Survey team. Antibody responses and correlates of protection in the general population after two doses of the ChAdOx1 or BNT162b2 vaccines. Nat Med. 2022 May;28(5):1072–1082. doi: 10.1038/s41591-022-01721-6.

30) Marra AR, Kobayashi T, Suzuki H, Alsuhaibani M, Tofaneto BM, Bariani LM, Auler MA, Salinas JL, Edmond MB, Doll M, Kutner JM, Pinho JRR, Rizzo LV, Miraglia JL, Schweizer ML. Short-term effectiveness of COVID-19 vaccines in immunocompromised patients: A systematic literature review and meta-analysis. J Infect. 2022 Mar;84(3):297–310. doi:10.1016/j.jinf.2021.12.035.

31) Embi PJ, Levy ME, Naleway AL, Patel P, Gaglani M, Natarajan K, Dascomb K, Ong TC, Klein NP, Liao IC, Grannis SJ, Han J, Stenehjem E, Dunne MM, Lewis N, Irving SA, Rao S, McEvoy C, Bozio CH, Murthy K, Dixon BE, Grisel N, Yang DH, Goddard K, Kharbanda AB, Reynolds S, Raiyani C, Fadel WF, Arndorfer J, Rowley EA, Fireman B, Ferdinands J, Valvi NR, Ball SW, Zerbo O, Griggs EP, Mitchell PK, Porter RM, Kiduko SA, Blanton L, Zhuang Y, Steffens A, Reese SE, Olson N, Williams J, Dickerson M, McMorrow M, Schrag SJ, Verani JR, Fry AM, Azziz-Baumgartner E, Barron MA, Thompson MG, DeSilva MB. Effectiveness of 2-Dose Vaccination with mRNA COVID-19 Vaccines Against COVID-19-Associated Hospitalizations Among Immunocompromised Adults – Nine States, January-September 2021. MMWR Morb Mortal Wkly Rep. 2021 Nov 5;70(44):1553–1559. doi: 10.15585/mmwr.mm7044e3.

32) Apostolidis SA, Kakara M, Painter MM, Goel RR, Mathew D, Lenzi K, Rezk A, Patterson KR, Espinoza DA, Kadri JC, Markowitz DM, E Markowitz C, Mexhitaj I, Jacobs D, Babb A, Betts MR, Prak ETL, Weiskopf D, Grifoni A, Lundgreen KA, Gouma S, Sette A, Bates P, Hensley SE, Greenplate AR, Wherry EJ, Li R, Bar-Or A. Cellular and humoral immune responses following SARS-CoV-2 mRNA vaccination in patients with multiple sclerosis on anti-CD20 therapy. Nat Med. 2021 Nov;27(11):1990–2001. doi:10.1038/s41591-021-01507-2.

33) Klineova S, Farber RS, DeAngelis T, Leung T, Smith T, Blanck R, Zhovtis-Ryerson L, Harel A. Vaccine-breakthrough SARS-CoV-2 infections in people with multiple sclerosis and related conditions: An observational study by the New York COVID-19 Neuro-Immunology Consortium (NYCNIC-2). Mult Scler. 2023 Jul;29(8):990–1000. doi:10.1177/13524585231185246.

34) Martins Figueiredo L, Carvalho E, Branco J, Carvalho Lourenço L, Santos L, Oliveira AM. Anti-tumor necrosis factor therapy is associated with attenuated humoral response to SARS-COV-2 vaccines in patients with inflammatory bowel disease. Vaccine. 2023 Jun 13;41(26):3862–3871. doi:10.1016/j.vaccine.2023.05.012.

35) Ruggeri EM, Nelli F, Fabbri A, Onorato A, Giannarelli D, Giron Berrios JR, Virtuoso A, Marrucci E, Mazzotta M, Schirripa M, Panichi V, Pessina G, Signorelli C, Chilelli MG, Primi F, Natoni F, Fazio S, Silvestri MA. Antineoplastic treatment class modulates COVID-19 mRNA-BNT162b2 vaccine immunogenicity in cancer patients: a secondary analysis of the prospective Vax-On study. ESMO Open. 2022 Feb;7(1):100350. doi:10.1016/j.esmoop.2021.100350.

36) Takeuchi M, Higa Y, Esaki A, Nabeshima Y, Nakazono A. Does reactogenicity after a second injection of the BNT162b2 vaccine predict spike IgG antibody levels in healthy Japanese subjects? PLoS One. 2021 Sep 20;16(9):e0257668. doi:10.1371/journal.pone.0257668.

37) Hwang YH, Song KH, Choi Y, Go S, Choi SJ, Jung J, Kang CK, Choe PG, Kim NJ, Park WB, Oh MD. Can reactogenicity predict immunogenicity after COVID-19 vaccination? Korean J Intern Med. 2021 Nov;36(6):1486–1491. doi:10.3904/kjim.2021.210.

38) Oyebanji OA, Wilson B, Keresztesy D, Carias L, Wilk D, Payne M, Aung H, Denis KS, Lam EC, Rowley CF, Berry SD, Cameron CM, Cameron MJ, Schmader KE, Balazs AB, King CL, Canaday DH, Gravenstein S. Does a lack of vaccine side effects correlate with reduced BNT162b2 mRNA vaccine response among healthcare workers and nursing home residents? Aging Clin Exp Res. 2021 Nov;33(11):3151–3160. doi: 10.1007/s40520-021-01987-9.

39) Hervé C, Laupèze B, Del Giudice G, Didierlaurent AM, Tavares Da Silva F. The how’s and what’s of vaccine reactogenicity. NPJ Vaccines. 2019 Sep 24;4:39. doi:10.1038/s41541-019-0132-6.

40) Mitchell TC, Casella CR. No pain no gain? Adjuvant effects of alum and monophosphoryl lipid A in pertussis and HPV vaccines. Curr Opin Immunol. 2017 Aug;47:17–25. doi:10.1016/j.coi.2017.06.009.

41) Burny W, Callegaro A, Bechtold V, Clement F, Delhaye S, Fissette L, Janssens M, Leroux-Roels G, Marchant A, van den Berg RA, Garçon N, van der Most R, Didierlaurent AM; ECR-002 Study Group. Different Adjuvants Induce Common Innate Pathways That Are Associated with Enhanced Adaptive Responses against a Model Antigen in Humans. Front Immunol. 2017 Aug 14;8:943. doi:10.3389/fimmu.2017.00943.

42) Collier AY, McMahan K, Yu J, Tostanoski LH, Aguayo R, Ansel J, Chandrashekar A, Patel S, Apraku Bondzie E, Sellers D, Barrett J, Sanborn O, Wan H, Chang A, Anioke T, Nkolola J, Bradshaw C, Jacob-Dolan C, Feldman J, Gebre M, Borducchi EN, Liu J, Schmidt AG, Suscovich T, Linde C, Alter G, Hacker MR, Barouch DH. Immunogenicity of COVID-19 mRNA Vaccines in Pregnant and Lactating Women. JAMA. 2021 Jun 15;325(23):2370–2380. doi:10.1001/jama.2021.7563.

43) Gray KJ, Bordt EA, Atyeo C, Deriso E, Akinwunmi B, Young N, Baez AM, Shook LL, Cvrk D, James K, De Guzman R, Brigida S, Diouf K, Goldfarb I, Bebell LM, Yonker LM, Fasano A, Rabi SA, Elovitz MA, Alter G, Edlow AG. Coronavirus disease 2019 vaccine response in pregnant and lactating women: a cohort study. Am J Obstet Gynecol. 2021 Sep;225(3):303.e1–303.e17. doi:10.1016/j.ajog.2021.03.023.

44) Atyeo CG, Shook LL, Brigida S, De Guzman RM, Demidkin S, Muir C, Akinwunmi B, Baez AM, Sheehan ML, McSweeney E, Burns MD, Nayak R, Kumar MK, Patel CD, Fialkowski A, Cvrk D, Goldfarb IT, Yonker LM, Fasano A, Balazs AB, Elovitz MA, Gray KJ, Alter G, Edlow AG. Maternal immune response and placental antibody transfer after COVID-19 vaccination across trimester and platforms. Nat Commun. 2022 Jun 28;13(1):3571. doi:10.1038/s41467-022-31169-8.

45) Herzberg J, Fischer B, Lindenkamp C, Becher H, Becker AK, Honarpisheh H, Guraya SY, Strate T, Knabbe C. Persistence of Immune Response in Health Care Workers After Two Doses BNT162b2 in a Longitudinal Observational Study. Front Immunol. 2022 Mar 4;13:839922. doi:10.3389/fimmu.2022.839922.

46) Malavazos AE, Basilico S, Iacobellis G, Milani V, Cardani R, Boniardi F, Dubini C, Prandoni I, Capitanio G, Renna LV, Boveri S, Rigolini R, Carrara M, Spuria G, Cuppone T, D’acquisto A, Carpinelli L, Sacchi M, Morricone L, Secchi F, Costa E, Menicanti L, Nisoli E, Carruba M, Ambrogi F, Corsi Romanelli MM. Antibody responses to BNT162b2 mRNA vaccine: Infection-naïve individuals with abdominal obesity warrant attention. Obesity (Silver Spring). 2022 Mar;30(3):606–613. doi:10.1002/oby.23353.

47) van der Klaauw AA, Horner EC, Pereyra-Gerber P, Agrawal U, Foster WS, Spencer S, Vergese B, Smith M, Henning E, Ramsay ID, Smith JA, Guillaume SM, Sharpe HJ, Hay IM, Thompson S, Innocentin S, Booth LH, Robertson C, McCowan C, Kerr S, Mulroney TE, O’Reilly MJ, Gurugama TP, Gurugama LP, Rust MA, Ferreira A, Ebrahimi S, Ceron-Gutierrez L, Scotucci J, Kronsteiner B, Dunachie SJ, Klenerman P; PITCH Consortium; Park AJ, Rubino F, Lamikanra AA, Stark H, Kingston N, Estcourt L, Harvala H, Roberts DJ, Doffinger R, Linterman MA, Matheson NJ, Sheikh A, Farooqi IS, Thaventhiran JED. Accelerated waning of the humoral response to COVID-19 vaccines in obesity. Nat Med. 2023 May;29(5):1146–1154. doi:10.1038/s41591-023-02343-2.

48) Ou X, Jiang J, Lin B, Liu Q, Lin W, Chen G, Wen J. Antibody responses to COVID-19 vaccination in people with obesity: A systematic review and meta-analysis. Influenza Other Respir Viruses. 2023 Jan;17(1):e13078. doi: 10.1111/irv.13078.

