## Supplementary Materials 1 for "Determinants of SARS-CoV-2 IgG response and decay in Canadian healthcare workers: a prospective cohort study"

1.1 Check list of immunological conditions
1.2 Question to elicit medications

1.3 Coding template for medications

1.4 Questions to elicit vaccine reactions

Supplementary Materials 1.1

C1 Do you have any of the following medical conditions?  Please check all that apply to you

- Rheumatoid arthritis (1)
- Ankylosing spondylitis (2)
- Psoriatic arthritis (3)
- Psoriasis (4)
- Inflammatory bowel disease (ulcerative colitis, Crohn’s disease) (5)
- Lupus (6)
- Other chronic autoimmune inflammatory disease? Please specify (7) __________________________________________________
- Other medical condition or treatment (except chemotherapy or organ transplant) that might affect an antibody response to vaccination? Please specify (8) __________________________________________________
- None of those listed (9)

Supplementary materials 1.2

C4 Since the start of pandemic, have you taken any medication that might **affect your response to a vaccination?** If taking medication and not sure if it might affect a response, please check not sure and give details in the grid below.

- Yes (1)
- No (4)
- Not sure (2)

*Display This Question:*

*If Since the start of pandemic, have you taken any medication that might affect your response to a v... = Yes*

*Or Since the start of pandemic, have you taken any medication that might affect your response to a v... = Not sure*

| 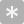 | 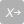 |
| --- | --- |

C4.1 Please give the details in the grid below

|  | When did you first start taking this Medication **MONTH** | When did you first start taking this Medication **YEAR** | For what conditions or symptoms are you taking this medication? | What tablets or other medications have you used? | How often do you take/use it? |
| --- | --- | --- | --- | --- | --- |
|  |  |  | (1) | (1) | (1) |

Supplementary materials 1.3

Medication reported by cohort study participants, by ICD-11 category

| **Medication category** | **Medication reported by study participants** |
| --- | --- |
| Methotrexate | Methotrexate |
| TNF inhibitors | Adalimumab, Certolizumab, Etanercept, Golimumab, Infliximab |
| Glucocorticoids | Hydrocortisone, Prednisone |
| Interleukin inhibitors | Ixekizumab, Secukinumab, Ustekinumab |
| Calcineurin inhibitors | Tacrolimus, Ciclosporin |
| Selective Immunosuppressants | Fingolimod, Upadacitinib, Ocrelizumab, Mycophenolic acid, Vedolizumab, Leflunomide, Ponesimod |
| Antineoplastic agents | Rituximab, Daratumumab, Cladribine, Dasatinib, Bortezomib |

1.4 Questions to elicit vaccine reactions

B4.3_2v Please mark on the line below **how** much were you bothered by side effects to the vaccine AFTER YOUR 2nd SHOT?

|  | Not at all   bothered | Very  bothered |
| --- | --- | --- |

| Fatigue (1) |
| --- |
| Myalgia (muscle pain) (21) |
| Arthralgia (joint pain) (22) |
| Headache (23) |
| Malaise (24) |
| Feeling feverish (25) |
| Chills (26) |
| Diarrhea/ loose stools (27) |
| Nausea/vomiting (28) |
| Skin reaction/rash (29) |
| Swollen glands (30) |
| Pain at injection site (31) |
| Redness at injection site (32) |
