## Supplementary Materials 2 for "Determinants of SARS-CoV-2 IgG response and decay in Canadian healthcare workers: a prospective cohort study"

Geometric mean anti-RBD IgG by calendar month of sample collection.

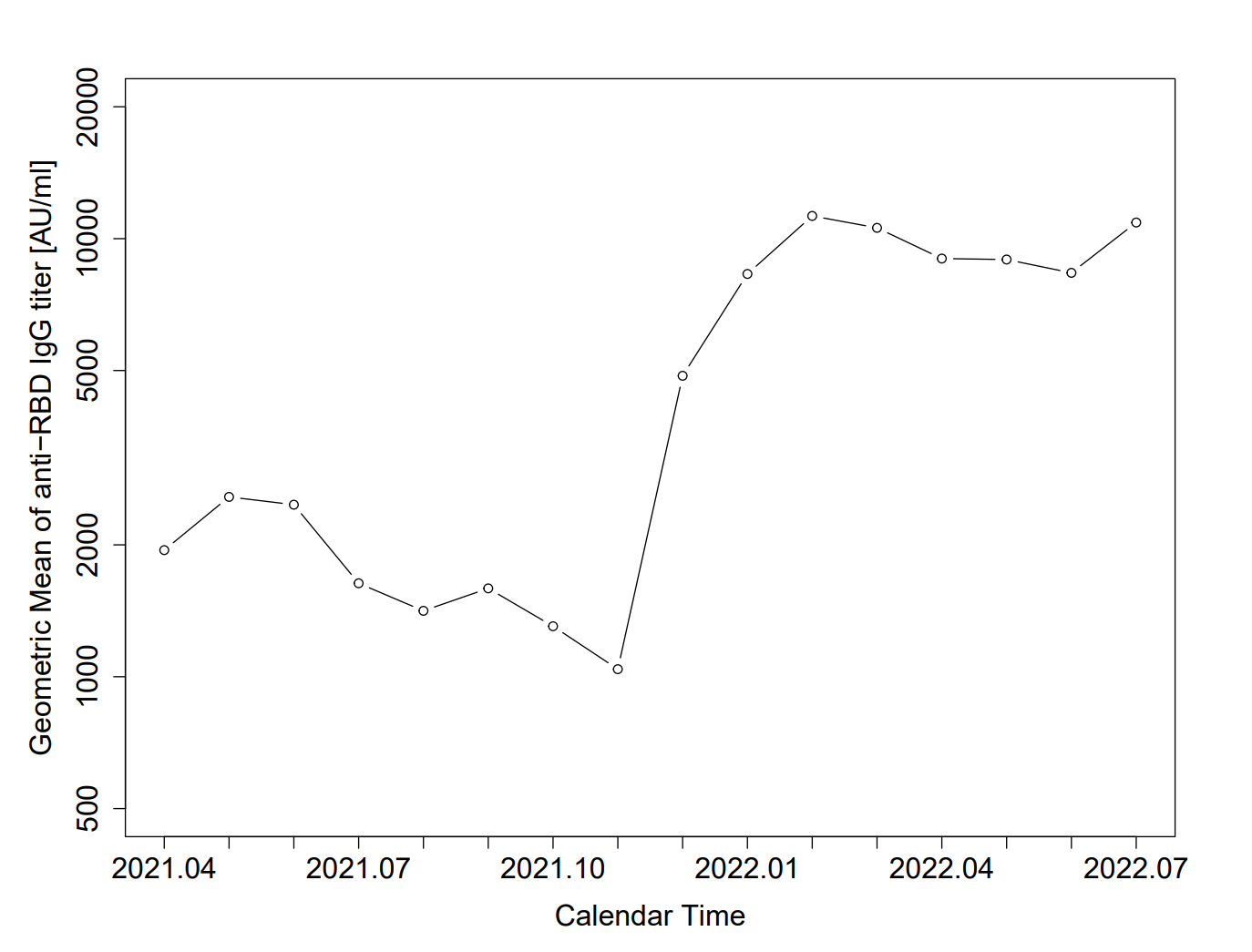
